# Association of Renin-Angiotensin-Aldosterone System Inhibition with Risk of COVID-19, Inflammation Level, Severity and Death in Patients With COVID-19: A Rapid Systematic Review and Meta-Analysis

**DOI:** 10.1101/2020.05.20.20108399

**Authors:** Xiao Liu, Chuyan Long, Qinmei Xiong, Chen Chen, Jianyong Ma, Yuhao Su, Kui Hong

**Affiliations:** Cardiology Department, the Second Affiliated Hospital of Nanchang University, Jiangxi, 330006, China; Jiangxi Key Laboratory of Molecular Medicine, Jiangxi, 330006, China

**Keywords:** COVID-19, ACEI/ARB, SARS-COV-2, pneumonia, infectious disease, lung, hypertension

## Abstract

**Background:** An association among the use of angiotensin-converting-enzyme(ACE) inhibitors and angiotensin-receptor blockers(ARBs) with the clinical outcomes of coronavirus disease 2019 (COVID-19) is unclear.

**Methods:** PubMed, EMBASE, and the preprint databases MedRxiv and BioRxiv were searched for relevant studies that assessed the association among inflammation level, application of ACEI/ARB, infection severity and death in patients with COVID-19. Odd risks(OR) and 95% confidence interval (CI) were combined using random-effects or fixed models depending on heterogeneity.

**Results:** Eleven studies were included with a total of 33,483 patients. Our review showed ACEI/ARB therapy might be associated with the reduced inflammatory factor (interleukin-6) and elevated level of immune cells(CD3, CD8). Meta-analysis showed no significant increase in the risk of COVID-19 infection(OR:0.95, 95%CI:0.89-1.05) in patients receiving ACEI/ARB therapy, and ACEI/ARB therapy was associated with a decreased risk of severe COVID-19 (OR:0.75, 95%CI: 0.59-0.96) and mortality (OR:0.52, 95%CI: 0.35-0.79). Subgroup analyses showed that, among the general population, application of ACEI/ARB therapy was associated with reduced risks of all-cause death(OR:0.31, 95%CI: 0.13-0.75), and the risk of severe COVID-19(OR:0.79, 95%CI: 0.60-1.05) infection and COVID-19 infection(OR:0.85, 95% CI: 0.66-1.08) were not increased. Among patients with hypertension, the use of an ACEI/ARB was associated with a lower severity of COVID-19(OR:0.73, 95%CI: 0.51-1.03) and lower mortality(OR:0.57, 95%CI: 0.37-0.87), without evidence of an increased risk of COVID-19 infection(OR:1.00, 95%CI: 0.90-1.12).

**Conclusion:** On the basis of the available evidence, this is the first meta-analysis showed that, in general population, the use of ACEI/ARB therapy was safe without an increased risk of COVID-19 infection and with a decreasing trend of severe COVID-19 infection and lower mortality. In patients with hypertension, the use of ACEI/ARB therapy should be encouraged, without increased risk of COVID-19 inflection, and better prognosis (a decreasing trends of severe COVID-19 and reduced all-cause death). Overall, ACEI/ARB therapy should be continued in patients who are at risk for, or have COVID-19, either in general population or hypertension patients. Our results need to be interpreted with caution considering the potential for residual confounders, and more well-designed studies that control the clinical confounders are necessary to confirm our findings.

## Introduction

The coronavirus disease 2019 (COVID-19) pandemic is becoming one of the most far-reaching public health crises in recorded history. At right time of writing this review, on April 29^th^, 2020, the number of infected persons worldwide has exceeded 3.01 million, with more than 200,000 reported deaths (http://2019ncov.chinacdc.cn/2019-nCoV/global.html). At present, there is no specifically targeted, effective treatment for patients with COVID-19. There is an urgent need, therefore, to determine how to alleviate the severe clinical symptoms and reduce the morbidity and mortality due to this disease. Severe acute respiratory syndrome coronavirus 2 (SARS-CoV-2) shares the same cell entry receptor as SARS-CoV, in which the viruses’ spike proteins bind to the host cell surface. Angiotensin converting enzyme 2 (ACE2) receptors, which are an essential regulator of renin–angiotensin–aldosterone system (RAAS) activity^1, 2^ The RAAS is a vital regulator of cardiovascular and renal function, including blood pressure, and which plays a vital role in regulating acute lung injury^3, 4^ The ACE2 level is significantly reduced in patients following SARS-CoV infection, resulting in RAAS system imbalance, and eventually caused severe acute lung injury. Angiotensin I converting enzyme inhibitors (ACEIs) and Angiotensin II receptor blockers (ARBs) are ACE2 receptor antagonists that can reduce the activity of the RAAS system and which have been widely used in the past several decades to treat cardiovascular diseases such as hypertension and hypertrophic cardiomyopathy.^3^ However, some animal experiments and clinical trials have shown that ACEIs potentially result in an increase in ACE2 receptors^5^. Some concern has therefore been expressed that the use of ACEIs/ARBs might increase the risk of COVID-19 infection after exposure to SARS-CoV-2 and result in a poor prognosis^6^. On April 12^th^, 2020, the European Society of Hypertension COVID-19 Task Force issued the following statement: the current evidence does not support that RAAS inhibitors aggravate the condition of COVID-19 patients^5^.

Accumulating evidence revealed that inflammatory cytokine storm and dysfunction of immune system largely contribute to the severe COVID-19, even cause death^7^. The latest evidence shows that peripheral blood interlekin-6 (IL-6) levels, a critical mediator of respiratory failure, shock, and multiorgan dysfunction, were significantly increased in COVID-19 patients who had used a RAAS inhibitor^8, 9^ Moreover, CD3 and CD8 cell counts were significantly reduced, suggesting that the level of inflammation in patients treated with ACEIs is relatively low^8^. Several retrospective studies have shown that the use of ACEIs/ARBs prior to COVID-19 infection could reduce the severity of COVID-19 and was associated with a better prognosis, especially in patients with both hypertension and COVID-19 ^10-12^ However, other studies have shown that there is no significant correlation between the use of ACEIs/ARBs and the severity and mortality of patients with COVID-19^10, 13-17^ Because the affection of RAAS inhibitors on the clinical prognosis of COVID-19 patients is unclear, in this article, we will review the current evidence and assess the clinical prognosis of COVID-19 patients with or without hypertension treated with ACEs/ARBs.

## Methods

This study was performed according to PRISMA guidelines (http://www.prisma-statement.org; **Table S1** Online Data Supplement)^18^.

### Literature Search

Two authors (L X. and Y.L) independently searched the PubMed and Embase databases for published articles and the preprint platforms medRxiv (https://www.medrxiv.org/) and bioRxiv (https://www.biorxiv.org/) (since many studies are available on these websites prior to publication, which allows for collection of the latest data) without language restrictions. Furthermore, the research references were traced and cross-checked. In the case of any discrepancy, it was resolved by consensus with the third author (H.K). The databases were searched for articles from February 1, 2019 to May 1, 2020 using the following search terms: ACE inhibitor, angiotensin II receptor blocker, angiotensin-converting enzyme inhibitor, 2019-novel coronavirus, SARS-CoV-2, COVID-19, and 2019-nCoV.

### Study Selection

Studies were considered eligible for inclusion if they 1) were designed as a randomized controlled trial, case-control study, or cohort study and 2) assessed the relationship between ACEI/ARB use and the level of inflammation, disease severity, and mortality in patients with COVID-19. If multiple studies used the same population, we selected the most recent publication. Certain publication types (e.g., reviews, editorials, letters, conference abstracts, and animal studies) or studies with insufficient data were excluded from this analysis.

### Data Extraction and Quality Assessment

Two authors (L X. and Y.L) independently extracted the study information and the basic characteristics of the articles using a standardized form, including the first author, year of publication, country, sample size, sex ratio, age, sample size, RAAS type, length of follow-up, adjustments for confounders and adjusted odds ratio (OR) with 95% confidence intervals (CIs). Two authors independently used modified Jada scores and Newcastle-Ottawa Scale (NOS)^19^ scores to evaluate the quality of the RCTs (randomized controlled trials) and observational studies, respectively. A Jada score >5 and an NOS score >7 were considered high quality scores^20^.

### Statistical Analyses

RevMan5.3 (Review Manager [RevMan], version 5.3, Cochrane Collaboration) software was used for statistical data processing, and the OR and 95% CIs were used to estimate the effect. Heterogeneity among the studies was analyzed using the *I*^2^ test with the following interpretation: low heterogeneity, defined as *I^2^*<50%; moderate heterogeneity, defined as *I^2^*=50% to 75%; and high heterogeneity, defined as *I^2^* >75%^21^. The meta-analysis was performed using the random effects model when the heterogeneity was more than 25%, otherwise, the fixed effects model was applied. If there were ACEI and ARB subgroups in the study, the combined OR and the 95% CIs were pooled using RevMan5.3 software. If the study did not report the OR directly, count data for outcomes were used to generate unadjusted ORs and 95% CIs. We also excluded reports with unadjusted ORs in the sensitivity analysis to assess the robustness of our results. A funnel plot was used to test for the presence of publication bias, and *P* value < 0.05 was considered statistically significant.

## Results

### Study Selection

The systematic search of the electronic databases identified 343 articles (PubMed=54, EMBASE=112, Medrxiv=132, ArXiv=45). After excluding duplicates and title/abstracts screened, 22 articles underwent a more detailed full-text assessment, after which a total of 11 articles with 33,483 patients were included^8, 10-17, 22, 23^ (**Figure 1**).

**Figure 1.**
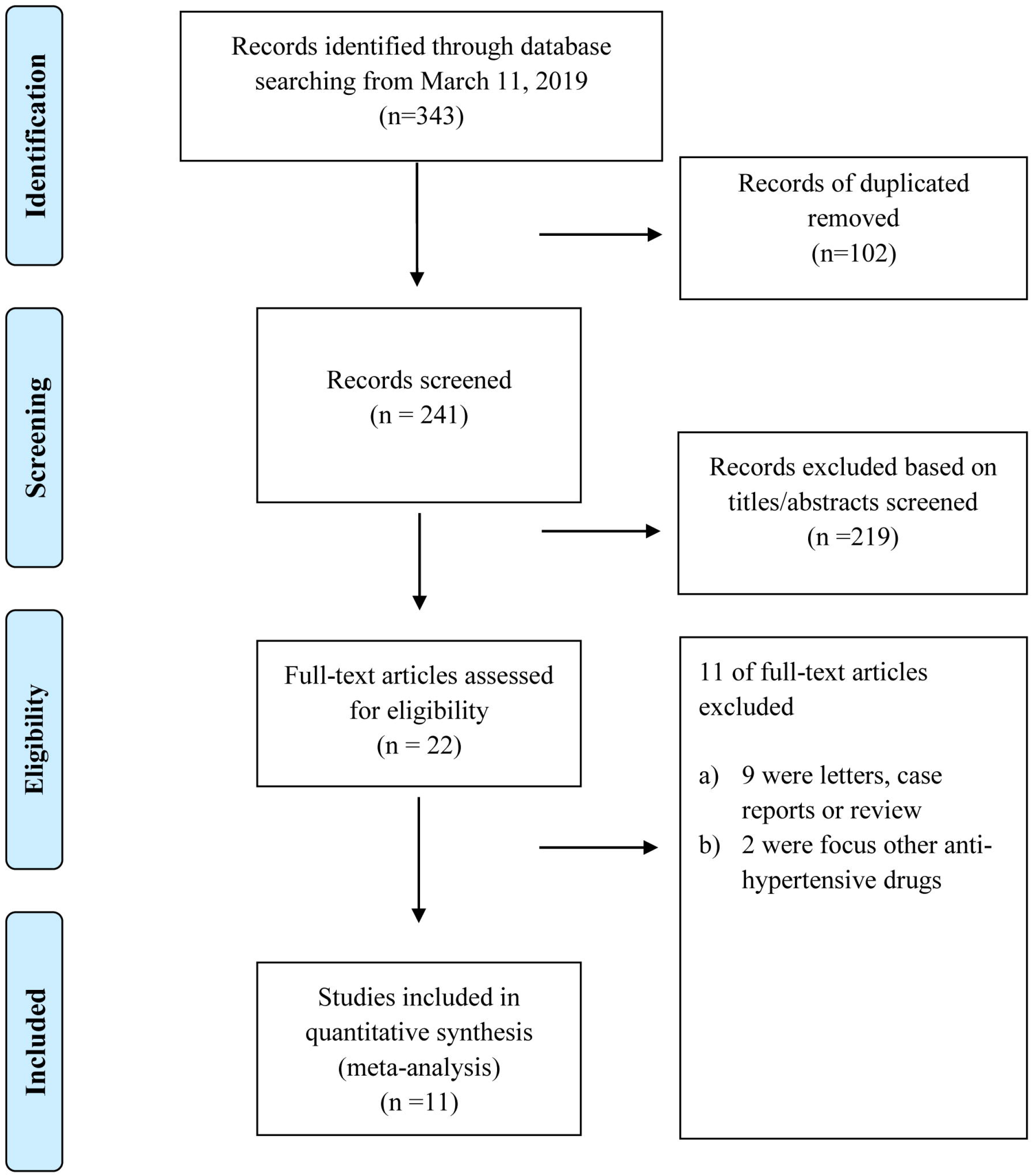
PRISMA Flow Diagram.

### Study Characteristics and Quality

**Table 1** shows the basic characteristics of the included studies. Overall, the sample sizes included in the articles ranged from 42 to 8,910, and ages ranged from 49 to 67 years old. Among the 11 studies, 7 were published^8, 10, 12, 15-17, 23^ and 4 were found on a preprint server^11, 14, 22^; 3 studies were based on the general population of COVID-19 patients ^11, 12, 15^ and 8 were based on patients with COVID-19 and hypertension^8, 10, 13, 14, 16, 17, 22, 23^. Four articles reported the effect of ACEIs/ARBs on the level of inflammation ^8, 10, 13, 17^, two studies assessed the risk of COVID-19 inflection^15, 16^, and all included studies evaluated the severity of disease or/and mortality^8, 10-17, 22, 23^. All included studies were observational studies. The NOS score of all of the observational studies was > 6, indicating that all of the studies were of high quality (**Table S2** Online Data Supplement).

### Inflammation Level

There were 4 articles that evaluated the relationship between ACEI/ARB and level of inflammation in patients with COVID-19^8, 13, 17, 22^, but since the data were not reported in the same unit across studies, they were not pooled; therefore, we conducted a systematic review instead (**Table 2**). Meng et al^8^. first assessed the impact of RAAS inhibitors on the level of inflammation in patients with COVID-19, and although they found no significant change in CRP levels from the peripheral blood in patients receiving ACEI/ARB treatment, IL-6 levels were significantly reduced, and the level of immune cells (CD3, CD8) counts were significantly increased.

In contrast, another study showed that there was no difference in IL-6 levels, while high-sensitivity C-reactive protein(CRP) test were significantly decreased^13^. A study that included 342 COVID-19 patients with hypertension who had taken ACEI/ARB drugs found that IL-6 levels were significantly reduced, while CRP level show no significantly different^17^. More recently, a multi-center study including 1,128 patients with hypertension and COVID-19 reported that the number of patients with high CRP values who had received ACEI/ARB therapy showed no significantly different than in the patients who had not received ACEI/ARB therapy^10^.

### Risk of Covid-19 Infection

Two^15, 16^ studies including a total of 10,629 patients reviewed the risk of COVID-19 infection, including one general population-based study and one study including patients with hypertension. There was no significant increase in the risk of COVID-19 infection in patients receiving ACEI/ARB therapy (OR=0.95, 95% CI: 0.89-1.02; *P*=0.14, *I^2^*=0%), with no significant heterogeneity (**Figure 2A**). Subgroup analysis based on population showed that the results did not change in the general population (OR=0.96, 95% CI: 0.89-1.05; *P*=0.45, *I^2^*=0%) or in the hypertensive population (OR=1.00, 95% CI: 0.90-1.12; *P*=1.00) (**Figure 3A)**.

**Figure 2.**
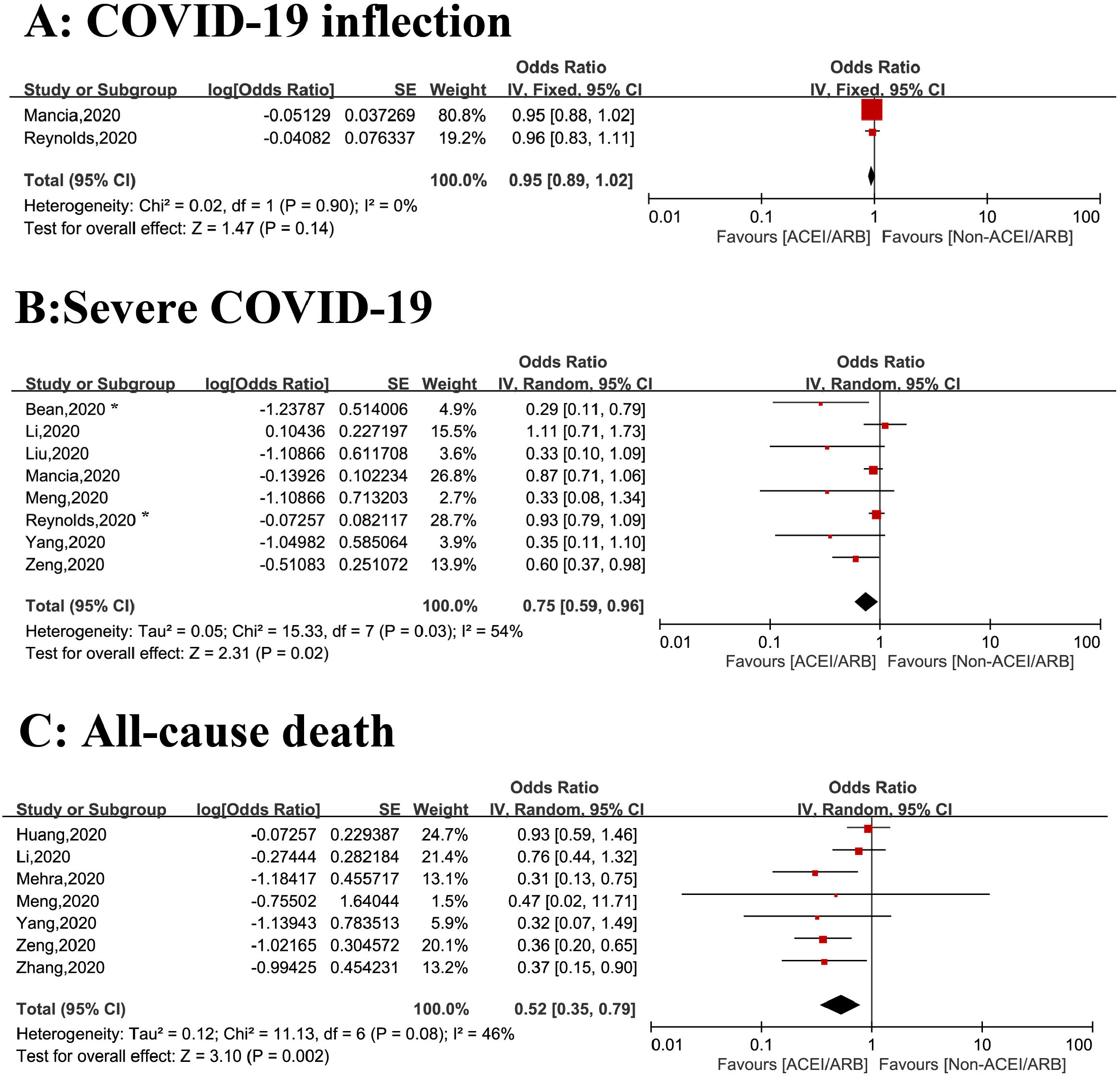
Summary of the associations between use of ACEI/ARB and clinical outcomes among patients with COVID-19. A. Risk of COVID-19 infection; B. Risk of severe COVID-19 infection; C. All-cause death. *severe COVID-19 or death

**Figure 3.**
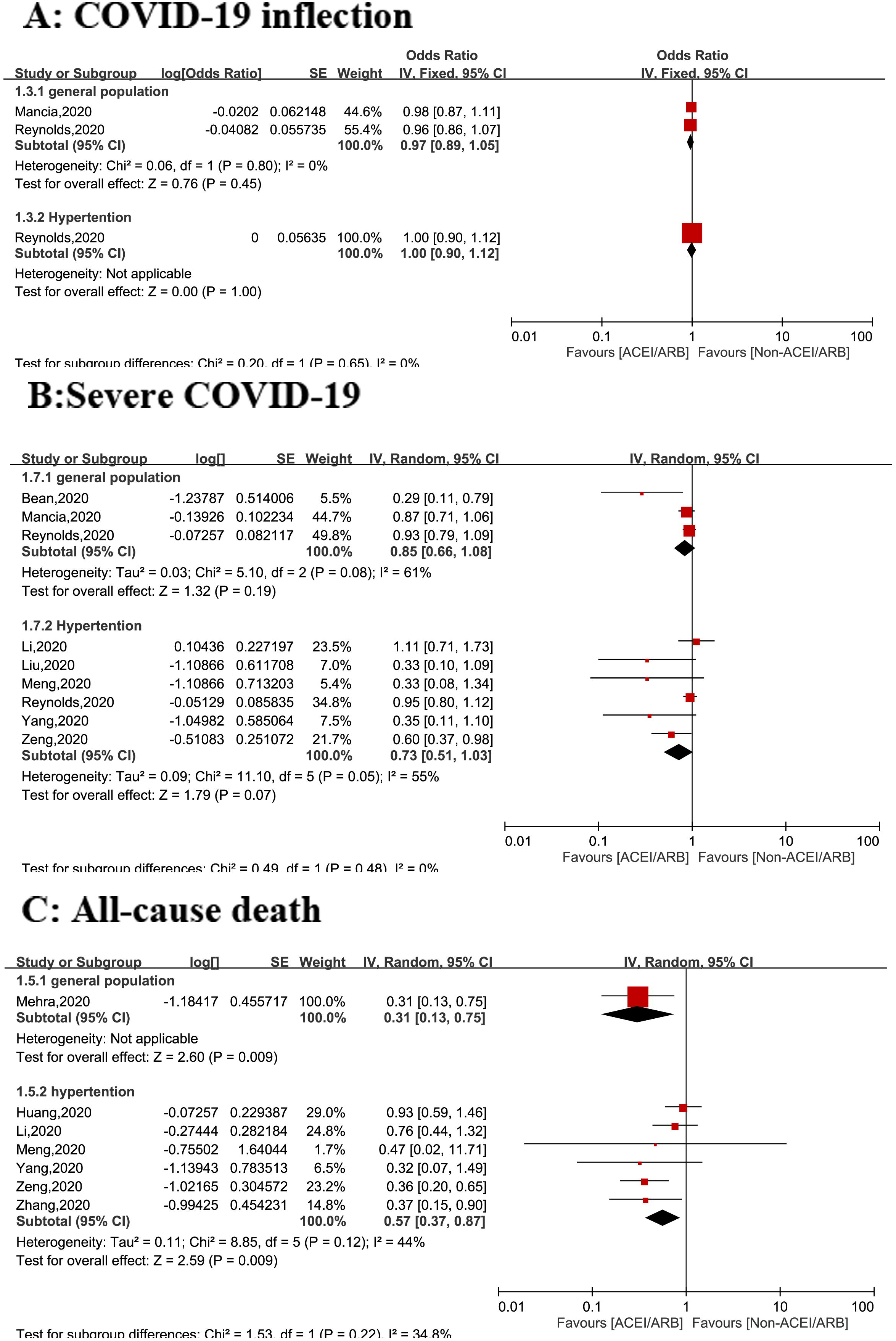
Subgroup analysis of the associations between use of ACEI/ARB and clinical outcomes among patients with COVID-19 stratified by general population and hypertensive population: A. Risk of COVID-19 infection; B. Risk of severe COVID-19 infection; C. All-cause death. *severe COVID-19 or death

### Risk of Severe COVID-19

Eight studies assessed the relationship between the use of ACEI/ARB therapy and severe COVID-19^8, 11, 13-17, 22^, with two reports in the general population^11,15^ and 6 reports in patients with hypertension^8, 13, 14, 16, 17, 22^ Compared with the non-ACEI/ARB group, the risk of severe COVID-19 infection decreased by 35% (OR=0.75, 95% CI: 0.59-0.96; *P*=0.02) in patients treated with an ACEI/ARB, with moderate heterogeneity (*I*^2^=54%) (**Figure 2B**). However, in the sensitivity analysis, the results were not statistically significant when the unadjusted studies were excluded (OR=0.85, 95% CI: 0.66-1.08; *P*=0.19).

Subgroup analysis showed there was no statistical significant association between the risk of severe COVID-19 infection and ACEI/ARB use either in the general population (OR=0.85, 95% CI: 0.66-1.08; *P*=0.19, *I*^2^=61%) or in hypertensive patients with (OR=0.73, 95% CI: 0.51-1.03; *P*=0.07, *I*^2^=55%) (**Figure 3B**).

### All-Cause Mortality

Seven studies reported all-cause mortality in 11,509 patients with COVID-19^8, 10, 12, 13, 17, 22, 23^, and ACEI/ARB therapy was associated with a decreased risk of all-cause mortality (OR: 0.52, 95% CI: 0.35-0.79; *P*=0.002) with modest of heterogeneity (*I*^2^=46%) (**Figure 2C**).There is no evidence of heterogeneity (*I*^2^=1%) when one^23^ study was excluded, with no change of the conclusion (OR: 0.46, 95% CI: 0.33-0.64; P<0.001). In the sensitivity analysis, the results were not statistically significant when the unadjusted studies were excluded (OR=0.34, 95% CI: 0.18-0.63; *P*=0.007, *I*^2^=0%).

Only one^12^ study reported all-cause mortality in COVID-19 patients in the general population, and which found that ACEI/ARB therapy significantly decreased the mortality rate of COVID-19 patients (OR: 0.31, 95% CI: 0.13-0.75; *P*=0.009). In hypertensive patients, the use of an ACEI/ARB was associated with a decreased risk of all-cause mortality (OR: 0.57, 95% CI: 0.37-0.87; *P*=0.009) (**Figure 3C**).

### Publication Bias

Publication bias was not detected in this study, as tests of publication bias are unsuitable when the number of included studies for each outcome is less than 10^24^.

## Discussion

This study was the first systemic assessment of ACEI/ARB therapy and the clinical prognosis of patients infected with COVID-19. We did not find an association between ACEI/ARB therapy and the increased risk of COVID-19 infection either in general population or in hypertensive patients. Moreover, we found that ACEI/ARB therapy could reduce the risk of severe COVID-19; in a subgroup analysis, RAAS inhibitors reduced the risk of severe COVID-19 by 15% and 27% in the general population and the hypertensive population, respectively, although this benefit did not achieve statistical significance. Finally, ACEI/ARB therapy reduced the rate of all-cause death in patients with COVID-19 by 48%, and in the subgroup analysis, use of an ACEI/ARB reduced the all-cause mortality by 69% and 43% for the general population and the hypertensive population, respectively.

Studies have shown that, similar to SARS-CoV, SARS-CoV-2 directly binds to the host cell ACE2 receptor which is highly expressed in human lung tissue, gastrointestinal tract, vascular endothelial cells, and arterial smooth muscle cells *in vivo^1, 25^*. Animal tests have shown that the expression of ACE2 receptors is significantly increased during ACEI/ARB therapy^5^, leading some scholars to worry that the use of RAAS system inhibitors may contribute to the spread of COVID-19 in the population^6^. On April 13^th^, 2020, the European Society of Hypertension COVID-19 Task Force updated their recommendations on the use of RAAS inhibitors in patients with COVID-19 and stated that the available evidence does not support a deleterious effect of RAAS blockers in COVID-19 infections^5^. Our research also determined that there is no significantly increased risk of infection with SARS-CoV-2 in patients receiving ACE/ARB therapy. There are several possible explanations for these findings. First, although some clinical studies have reported that RAAS inhibitors might increase the expression of ACE2 receptors, these results are inconsistent^5^ and most are small studies with small sample sizes^26-28^. Furthermore, these studies detected circulating ACE2 or soluble ACE2 receptor levels in the urine, and the level of soluble ACE2 receptors may not accurately reflect the true level of ACE2 receptors in the organs and tissues^26-28^. One study assessed the expression of ACE2 receptors in the duodenal and ileal tissues of 21 patients with RAAS inhibition; the researchers found that the expression of ACE2 receptor mRNA and protein increased significantly in the brush border of the small intestinal epithelial cells^29^. However, we know that SARS-CoV-2 is mainly transmitted through the respiratory tract ACE2 receptors^1, 30^, and to date, there have been no reports published on the expression of ACE2 receptors in lung tissue after ACEI/ARB treatment, which is an important direction for future study.

Our study found that RAAS inhibitors reduced the risk of severe COVID-19 infection by 47%, which might be related to the previously confirmed anti-inflammatory effects of RAAS inhibitors^4^. ACEIs and AT1R inhibitors are commonly used RAAS system inhibitors that have been widely used in the treatment of hypertension, diabetic nephropathy, and congestive heart failure^3^. Lot of studies have shown that RAAS system is an important target for the treatment of acute lung injury^31^. For example, a retrospective analysis showed that ACEIs and AT1R inhibitors can also reduce the incidence of radiation pneumonitis^32^. In patients with severe COVID-19, the levels of many pro-inflammatory factors (e.g., IL-6, IL-2, and TNF-α) were significantly elevated, and levels of regulatory T cells decreased significantly^7^. A non-randomized controlled clinical trial showed that the IL-6 inhibitor tocilizumab could significantly reduce oxygen consumption, imaging abnormalities, and clinical prognosis in patients with COVID-19^33^. Moreover, in patients receiving ACEI/ARB therapy, IL-6 and CRP levels were significantly decreased, and the level of CD3 andCD8 increased significantly^8^. Overall, this evidence suggests that ACEI/ARB therapy might reduce lung injury and infection severity by downregulating inflammation levels in patients with COVID-19.

Although we found use of ACEI/ARB was associated with the decreased risk of severe COVID-19 in the overall analysis, this benefit did not reach statistical significance in the sensitive analysis (with excluding unadjusted studies).Furthermore, subgroup analysis showed the benefit was also not statistically significant either the general population or in patients with hypertension. The possible reason for this result may be related to the limited sample size and existence of many confounding factors in the baseline characteristics such as cardiovascular disease, and the use of drugs may affect these results. Age has been proven to be a strong risk factor for both infection with COVID-19 and a higher severity of COVID-19 infection^12^. Additionally, population-based studies have estimated that the prevalence of hypertension range 9.5%-31.2% in COVID-19 patients, and many studies have shown that hypertension can also significantly increase the severity of COVID-19^34^. Therefore, more studies with large sample sizes with adjusted for these potential confounders are needed to clarify the impact of ACEI/ARB use on the severity of COVID-19 in both the general population and in hypertensive patients. Recently, one multi-center study that included 1,128 adult patients with hypertension also showed that ACEI/ARB use reduced the risk of septic shock by 62% without reducing the risk of other serious complications such as acute respiratory distress syndrome and disseminated intravascular coagulation after matching baseline factors, suggesting that ACEI/ARB therapy led to a different type of severity of COVID-19^10^. Therefore, the effect of RAAS inhibitors on the severity of COVID-19 remains controversial and requires further study.

In addition, we found that ACEI/ARB therapy reduced the risk of mortality by 47%, and in the subgroup analysis, this mortality benefit remained significant for both in the general population and in hypertensive patients. These results remained consistent in the sensitivity analysis, which verified the robustness of our study. The number of studies included in the subgroup of general population, however, was relatively limited (N=1)^12^, and the case-control study design increases bias (e.g., recall bias). Finally, we observed that 26.3% of the population carriedhypertension^12^, and studies have shown that COVID-19 patients with hypertension receiving ACEI/ARB therapy have lower mortality compared with those non-hypertensive individuals with COVID-19. It is therefore unclear whether the benefit was attributable to the high proportion of patients with hypertension. Moreover, the study only adjusted for age and sex and did not correct for enough of the potential confounding factors, such as drug use and hypertension. Therefore, the benefits of RAAS inhibition in the general population with a lower prevalence of hypertension still needs to be confirmed in future research.

### Study Limitations

Our research has several limitations. First, all of the included articles were observational and therefore cannot confirm the cause-and-effect relationship between ACEI/ARB therapy and the clinical prognosis of patients with COVID-19; a large-scale RCT is needed to confirm our results. Second, coexisting conditions such as hypertension have been shown a key prognostic determinant (e.g. severity and mortality) in patients with COVID-19. The guidelines recommend that patients with hypertension and COVID-19 continue their ACEI/ARB therapy, and our study reinforced this recommendation and further showed that the use of ACEI/ARB therapy might be associated with better clinical outcomes in the general population with COVID-19 and in hypertensive patients with COVID-19. However, the benefit of RAAS inhibitors in non-hypertensive patients might differ from those with hypertensive patients. Due to data limitations, we cannot analyze the severity and clinical prognosis of ACEI/ARB therapy in patients with COVID-19 without hypertension. Third, studies have shown that

ACEIs and ARBs may play different roles in patients with COVID-19, due to limited data, we were unable to perform subgroup analyses of ACEIs and ARBs. Fourth, some characteristic clinical values (e.g., drug variables) were missing. For example, the specific details of RAAS inhibitors were lacking in all studies, which might have impact on our results. Fifth, considering that all of the included studies were retrospective, the introduction of recall bias was inevitable and may affect the reliability of the conclusions. Sixth, although we did not register the protocol of this meta-analysis in the PROSPERO database, no relevant protocols of this topic were found in this database at the writing of this review.

## Conclusions

On the basis of the available evidence, we found that the use of ACEI/ARB therapy was not associated with an increased risk of COVID-19 and that there was a decreased trend between ACEI/ARB use and severe COVID-19 infection in both the general population and in patients with hypertension, although this association was not statistically significant. The risk of all-cause death was decreased with ACEI/ARB therapy in both the general population and in patients with hypertension. Overall, RAAS inhibitors should be continued use in patients who are at risk for, are being evaluated for, or have COVID-19. Our results need to be interpreted with caution considering the potential for residual confounders, and more well-designed studies that control clinical confounders are necessary to confirm our foundlings.

## Data Availability

All data generated or analyzed during this study are included in this published article [and its supplementary information files].

## Acknowledgements

K.H. was responsible for the entire project and revised the draft. X.L. and Y.L. performed the systematic literature review and drafted the first version of the manuscript. M.X., H.S, C.C and Y.M reviewed, interpreted, and checked data. All authors took part in the interpretation of the results and prepared the final version of the manuscript.

## Nonstandard Abbreviations and Acronyms

COVID-19: Coronavirus disease 2019
OR: Odd risk
CI: Confidence interval
ACE2: angiotensin-converting enzyme 2
ACEI: ACE inhibitors
ARB: ACE angiotensin–receptor blockers
NOS: Newcastle-Ottawa Scale score
RAAS: renin–angiotensin–aldosterone system
SARS: Severe acute respiratory syndrome.

